# Discovery of Ancestry-specific Variants Associated with Clopidogrel Response among Caribbean Hispanics

**DOI:** 10.1101/2023.09.29.23296372

**Authors:** Guang Yang, Pablo González, Mariangeli Moneró, Kelvin Carrasquillo, Jessicca Y. Renta, Dagmar F. Hernandez-Suarez, Mariana R. Botton, Kyle Melin, Stuart A. Scott, Gualberto Ruaño, Abiel Roche-Lima, Cristina Alarcon, Marylyn D. Ritchie, Minoli A. Perera, Jorge Duconge

## Abstract

**Background:** High on-treatment platelet reactivity (HTPR) with clopidogrel is predictive of ischemic events in adults with coronary artery disease. Despite strong data suggesting HTPR varies with ethnicity, including clinical and genetic variables, no genome-wide association study (GWAS) of clopidogrel response has been performed among Caribbean Hispanics. This study aimed to identify genetic predictors of HTPR in a cohort of Caribbean Hispanic cardiovascular patients from Puerto Rico.

**Methods:** Local Ancestry inference (LAI) and traditional GWASs were performed on a cohort of 511 clopidogrel-treated patients, stratified based on their P2Y12 reaction units (PRU) into responders and non-responders (HTPR).

**Results:** The LAI GWAS identified variants within the *CYP2C19* region associated with HTPR, predominantly driven by individuals of European ancestry and absent in those with native ancestry. Incorporating local ancestry adjustment notably enhanced our ability to detect associations. While no loci reached traditional GWAS significance, three variants showed suggestive significance at chromosomes 3, 14 and 22 (*OSBPL10* rs1376606, *DERL3* rs5030613, and *RGS6* rs9323567). In addition, a variant in the *UNC5C* gene on chromosome 4 was associated with an increased risk of HTPR. These findings were not identified in other cohorts, highlighting the unique genetic landscape of Caribbean Hispanics.

**Conclusion:** This is the first GWAS of clopidogrel response in Hispanics, confirming the relevance of the *CYP2C19* cluster, particularly among those with European ancestry, and also identifying novel markers in a diverse patient population. Further studies are warranted to replicate our findings in other diverse cohorts and meta-analyses.

## Introduction

Clopidogrel (Plavix®) is a P2Y12 receptor inhibitor that is widely recommended for secondary prevention of coronary artery disease (CAD) to reduce major adverse cardiovascular and cerebrovascular events (MACCEs) among patients with acute coronary syndrome (ACS) or stable CAD undergoing percutaneous coronary interventions (PCI). Despite its proven effectiveness as part of dual antiplatelet therapy (DAPT), some patients do not attain adequate antiplatelet effects.^1,2^ Studies have shown that certain genetic variants in clinically relevant pharmacogenes (e.g., *CYP2C19*\*2) are significantly associated with a diminished response to clopidogrel.^2,3^ Importantly, a meta-analysis of 9 clinical trials and 9,685 patients with ACS showed that carriers of one or two loss of function (LOF) *CYP2C19* alleles had 1.6-fold greater risk for MACCEs and 2.8-fold increased risk of stent thrombosis compared to non-carriers.^4^ Moreover, carrying at least one LOF *CYP2C19* allele resulted in a higher rate of MACCEs when compared to patients with a wild-type genotype (i.e., 12.1% vs. 8.0%).^5^ However, it is estimated that *CYP2C19* accounts for only ∼12% of the observed variability in clopidogrel response.^1^ In addition, approximately 22% of poor responders to clopidogrel do not carry any known reduced-function *CYP2C19* allele, suggesting that novel genetic markers or overlooked alleles for clopidogrel response prediction remain to be discovered. A prior genome-wide association study (GWAS) conducted by the International Clopidogrel Pharmacogenomics Consortium (ICPC) confirmed the significance of *CYP2C19*\*2 in clopidogrel resistance. It also identified novel genetic loci associated with risk of MACCEs in individuals with CAD, PCI, and ACS.^6^ However, it is important to note that the majority of these studies primarily involved individuals of European ancestry, with very limited representation from Hispanics and other minorities populations.^7^

Although routine implementation of *CYP2C19* testing to guide antiplatelet therapy selection is not currently standard of care, in part due to a paucity of large randomized controlled trials evaluating this approach, the recent American College of Cardiology Foundation/American Heart Association (ACC/AHA) ACS guidelines noted that genetic testing might be considered on a case-by-case basis to identify whether a high-risk patient is predisposed to inadequate platelet inhibition with clopidogrel.^8^ Moreover, the Clinical Pharmacogenetics Implementation Consortium (CPIC) recommends an alternative antiplatelet therapy (if not contraindicated) when *CYP2C19* genotype status is known for carriers of *CYP2C19* risk alleles, particularly for those with a poor metabolizer (PM) phenotype.^9^ A *CYP2C19* genotype-guided strategy for optimal selection of antiplatelet therapy has been proven beneficial in several clinical studies; however, none of these trials were performed with Caribbean Hispanic patients.^7,10–12^

To date, a substantial knowledge gap exists in our understanding of clopidogrel pharmacogenomics among Caribbean Hispanics, as they make up less than 1% of participants in previous studies. This lack of representation tends to exacerbate already existing healthcare disparities.^13–14^ Our study aims to address this gap by conducting a novel GWAS of residual on-treatment platelet reactivity in Caribbean Hispanic patients to identify genetic determinants of clopidogrel responsiveness in this underrepresented population. Furthermore, our study distinguishes itself as the first to incorporate local ancestry information into the genetic analysis of clopidogrel response, adding a novel dimension to our understanding of this critical pharmacogenomic trait.

## Methods

This was a multicenter, cross-sectional, clinical protocol to perform an unbiased pharmacogenomic association analysis of clopidogrel response in Caribbean Hispanics. Five hundred and eleven patients of Hispanic descent who reside in the Commonwealth of Puerto Rico and received either a 600 or 300 mg loading dose and a daily 75 mg maintenance dose of clopidogrel (i.e., alone or as a component of dual antiplatelet therapy (DAPT)) were recruited into the study. Enrollment occurred between January 2018 and June 2020 for patients who were prescribed clopidogrel after either PCI with ACS or for stable CAD with elective stenting. All patients received a daily maintenance dose of clopidogrel for a minimum of 7 days prior to enrollment. This study received Institutional Review Board (IRB) approval (#A4070417) and adhered to the study protocol (NCT03419325). **Figure *S1*** shows the number of participants who were screened, enrolled, and subsequently completed the study from seven different medical facilities across the island. The study used minimal inclusion and exclusion criteria (see ***Table S1***) to recruit a diverse population representative of real-world clinical practice, irrespective of platelet reactivity status or *CYP2C19* polymorphisms. Medication adherence was evaluated through self-reporting and record reviews, following a standardized methodology as previously described.^15^

Blood samples (20 mL) for genetic testing and rapid *ex vivo* residual platelet function analysis were collected at a single time point on the day of recruitment while under clopidogrel maintenance daily dosing of at least 7 days. Blood was collected into EDTA and sodium citrate (3.2%) tubes. Platelet reactivity was measured using the VerifyNow P2Y12 assay (Accumetrics, San Diego, CA, USA), following the manufacturer’s instructions (i.e., assessed within 4 hours after blood withdrawal but discarding the first volume of 3 mL of blood). The results were expressed as P2Y12 reaction units (PRU) and used to determine inhibition of platelets by clopidogrel in each participant. High on-treatment platelet reactivity (HTPR) was defined as PRU ≥230, which indicated poor clopidogrel response.^16–17^

### Genotyping and imputation

Genomic DNA was extracted using standard methods (Qiagen, CA, USA). Genotyping was performed using the HiScan® system with the whole-genome Infinium® Multi-Ethnic Hispanic AMR/AFR MEGA BeadChip array according to manufacturer instructions (Illumina, San Diego, CA, USA). Standard quality control (QC) procedures were implemented, which included: removal of single nucleotide polymorphisms (SNPs) with excessive missingness (> 5%) and individuals with high missing genotype (> 5%) or high heterozygosity rates (three standard deviations [SD] away from the mean rate). Hardy-Weinberg equilibrium (HWE) was calculated for all genotyped SNPs. Any SNPs that deviated from HWE were flagged but not removed. Gender misspecification was checked using X chromosome zygosity. Individuals who did not match known sample data were excluded. Identity-by-descent (IBD) check was performed to identify sample duplicates, contaminated samples, and cryptic relationships. For each pair of samples with estimated IBD coefficients greater than 0.185, only the sample with the highest call rate was retained. Only SNPs with a minor allele frequency (MAF) greater than 5% were included in our analyses. We then imputed the QCed genotype data using the TOPMed Imputation server (https://imputation.biodatacatalyst.nhlbi.nih.gov/#!), which were performed following standard procedures described elsewhere.^18^

### Genome-wide association studies (GWAS) analyses

Covariate frequencies (e.g., means ± [SDs]) and statistics for the study population as well as measures of HWE and linkage disequilibrium (LD) metrics were calculated using R and PLINK v1.9. After QC, 12,343,367 genotyped and imputed variants were evaluated for association. For each analysis, Manhattan, and quantile-quantile (Q-Q) plots were generated to visualize results. GWAS regression analyses using an additive genetic model were adjusted for relevant covariates (e.g., age, sex, body mass index (BMI), diabetes) and the first two principal components (PCs). SNPs with *p*-value ≤ 5·10^-8^ were considered as genome-wide significant, whereas SNPs with *p*-value ≤ 10^-6^ were considered as suggestive association. LocusZoom.js v0.12 was used to capture and visualize regions of interest.^19^ All statistical analyses were performed using PLINK v1.9. A separate multivariable logistic regression GWAS was performed to estimate the odds ratio and 95% CI for HTPR (high on therapy PRU, defined as ≥ 230) adjusted for age, sex, diabetes, BMI, PC1 and PC2 using PLINK v1.9.

### Population and Local Specific Ancestry Estimations

Global and local ancestry for each individual level were estimated to adjust the association analysis by admixture. Based on the history of admixture in the Caribbean Hispanic population, three different ancestral populations were utilized as reference panels for ancestry estimation. Genotype data from a total of 107 individuals of European ancestry (IBS: Iberian populations in Spain) and 61 of African ancestry (YRI: Yoruba in Ibadan, Nigeria) from the 1,000 Genomes project^20^ Phase 3, and 103 Native Mesoamerican and South American individuals from the Human Genome Diversity Project/*Centre d’Etude du Polymorphism* (HGDP-CEPH) database (AMR: Native American populations distributed as follows: Maya from southern Mexico; Colombian; Karitiana from western Amazon, Brazil; Surui from Mato-Grosso, Brazil; and Pima from central/southern Arizona and northwestern Mexico) were used to infer ancestry proportions and local ancestry.^21^

The reference panels were combined into a single VCF file that included the study cohort data using BCFtools v1.9^22^ and VCFtools v0.1.13^23^, for pre-processing and combining references with study samples. We used PLINK v1.9^24^ software to convert the combined VCF file into a PLINK binary file (BED/BIM/FAM), which was used as input for ADMIXTUREv1.3.0^25^ and RFMixv2^26^ software. For quality assurance and control, we performed a multidimensional PC analysis using PLINK to observe clustering between references and study data (Figure 1c). We used ADMIXTURE to estimate proportions of ancestry using a K=3, considering our three reference populations (YRI, IBS, and AMR). We used RFMix with a window size of 0.2cM, number of generations equal to 8 and the number of trees to generate per Random Forest set to 100.

**Figure 1.**
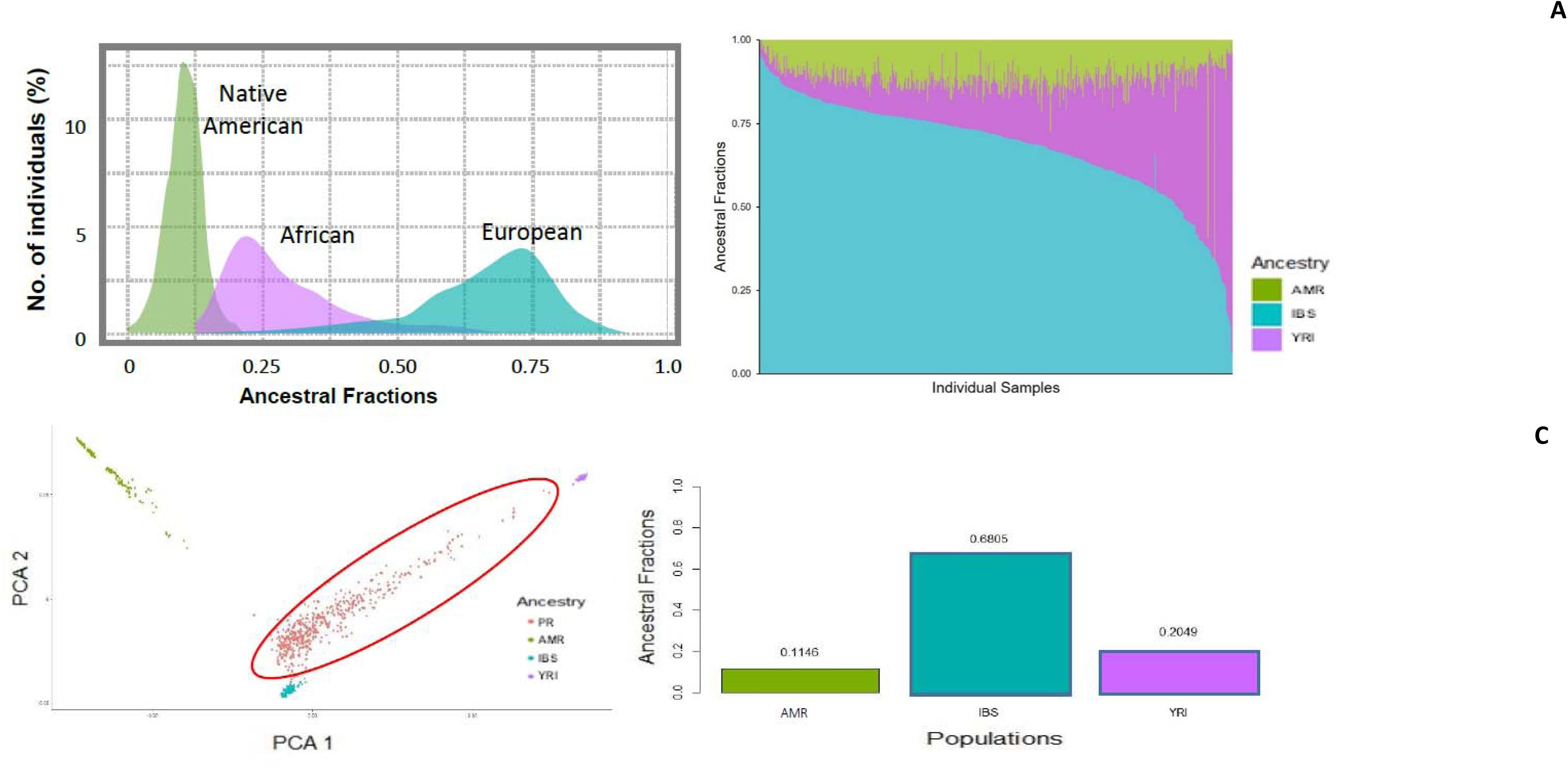
Genetic Admixture in Caribbean Hispanics. A) Density plots of ancestral proportions with green representing Native American (AMR: 103 Native Mesoamerican and South American individuals from the Human Genome Diversity Project/ *Centre d’Etude du Polymorphism* database (HGDP-CEPH); representing Maya and Pima in Mexico, Colombian in Colombia, Karitiana and Surui in Brazil), purple representing African (YRI: 61 Yoruba in Ibadan, Nigeria from 1,000 Genomes project), and cyan representing European (IBS: 107 Iberian populations in Spain from 1,000 Genomes project) ancestries, respectively. ^20–21^ There are different proportions of African and European contributions to the overall genetic background of each participant. Native American curve suggests this ancestral component is less variable among participants, but also smaller in terms of global proportion. B) The average level of individual ancestral proportions (fractions) per parental population in the study cohort. Each column represents an individual in the study cohort. C) Principal component analysis (PCA) in each participant of the study (PR: Caribbean Hispanics from Puerto Rico, in red color) along with the European, African, and Native American reference populations. D) Bar chart of the average ancestral proportions across the cohort. **Note:** when merged with non-overlapping SNPs typed previously in all individuals from our study cohort (PR), the resulting data for admixture analysis at both the population and regional (continental) level consist of over 30 million SNPs.

### Local ancestry inferences (LAI) adjusted GWAS

The LAI GWAS was performed to improve the power to detect association over standard GWAS using the Tractor software package^27^ adjusted for age, sex, diabetes, BMI, PC1, and PC2. Briefly, the RFMix v2 ancestry calls were converted to Tractor format, which includes genotype dosage and haplotype count information for African, Native American, and European ancestry at each SNP position. The Tractor local ancestry GWAS was then performed using PLINK. This analysis allowed us to use the LAI at each SNPs position as a SNP-based covariate in the GWAS for this admixed cohort. All clinical covariates that showed association to PRU or HTPR were included as covariates in the analysis. The deconvoluted model within TRACTOR was used in this analysis.^27^ Meta-analyses on the deconvoluted AFR, AMR, and EUR summary statistics was conducted using METAL using a random-effect model. We pre-specified SNPs at *p*-value ≤5·10^-8^ as significant, and those at *p*-value ≤10^-6^ as suggestive.

### Replication cohorts

The replication cohorts consisted of 167 African Americans and 200 Brazilians on maintenance doses of clopidogrel enrolled from 5 hospital system in Chicago and Washington DC (University of Chicago Medical Center, University of Illinois and Northwestern Memorial Hospital, George Washington University Hospital and Medical Faculty Associates, and the Washington DC VA Medical Center) through the African American Cardiovascular Pharmacogenomics Consortium (ACCOuNT)^28^, *Hospital de Clínicas de Porto Alegre*, and biorepository of the Universidade Federal do Rio Grande do Sul, in Porto Alegre, Brazil; these samples were independent from those used in the discovery cohort. All patients provided written, informed consent for study participation and were genotyped as described above via the Infinium® Multi-Ethnic MEGA BeadChip array. Available clinical data from the replication data set included sex, age, BMI, PRU measures, concomitant medications, type 2 diabetes, and indication for therapy. HTPR was pre-specified as PRU ≥ 230 on clopidogrel therapy as previously described. LAIs of each subject were estimated with RFMix v2^26^ as described above but using YRI and CEU samples from 1,000 Genomes phase 3^20^ as the reference populations (ACCOuNT cohort only). For the replication models, we selected the same clinical covariates used in the discovery models. Both PRUs and HTPR were modeled using significant covariates and the top hits by the linear and logistic regression model functions in PLINK v1.9. Because we tested only the three SNPs that were significant by regular GWAS in our discovery cohort when using a linear model (PRU) and the one identified after performing logistic regression (HTPR), as well as the top two SNPs in LAI GWAS, we did not correct for multiple hypothesis testing (i.e., *p*-value ≤0.05 significance was used instead). In addition, we used available data from the GWAS conducted in the ICPC cohort for external validation. A detailed description of the ICPC cohort and results can be found elsewhere.^6^

## Results

We recruited and analyzed a cohort of 511 Puerto Rican patients on maintenance dose of clopidogrel (75mg/day). **Table 1** describes the baseline characteristics of the participants from the study population with complete genetic profiles and clinical data. The average age of all participants in this study was ∼68 years old, with 45% identified as female. Most were middle-aged with a high prevalence of conventional risk factors (i.e., high BMI (28.4 kg/m^2^), 83.8% hypertension, 71.9% hypercholesterolemia/dyslipidemias, and 54.8% type-2 diabetes). Furthermore, 20% of participants were on proton pump inhibitors (PPI, mainly pantoprazole); whereas statins and calcium channel blockers (CCB) were prescribed in 79.1% and 26.8% of patients, respectively. Patients who were taking aspirin administered as part of DAPT represented 63.3% of the total cohort. Among 72.6% of participants, clopidogrel was given for stable CAD or ACS. About 18% of participants received a loading dose of clopidogrel (300 or 600 mg) on the scheduled date for PCI, while 78% were already receiving maintenance doses of clopidogrel 75mg/daily by the time they underwent the intervention and, therefore, a loading dose was not given. Since no differences in outcomes or baseline characteristics existed among patients with different dose schemes, they were combined for further analyses. Cases tended to be women, whose proportion significantly exceeded that of controls (59.8% vs. 40.6%; p< 0.001). Cases also showed a higher proportion of patients diagnosed with type-2 diabetes (63% vs. 52%; p=0.034), but less smokers (p=0.025). The cases also had a higher BMI on average (p=0.002).

**Table 1.**
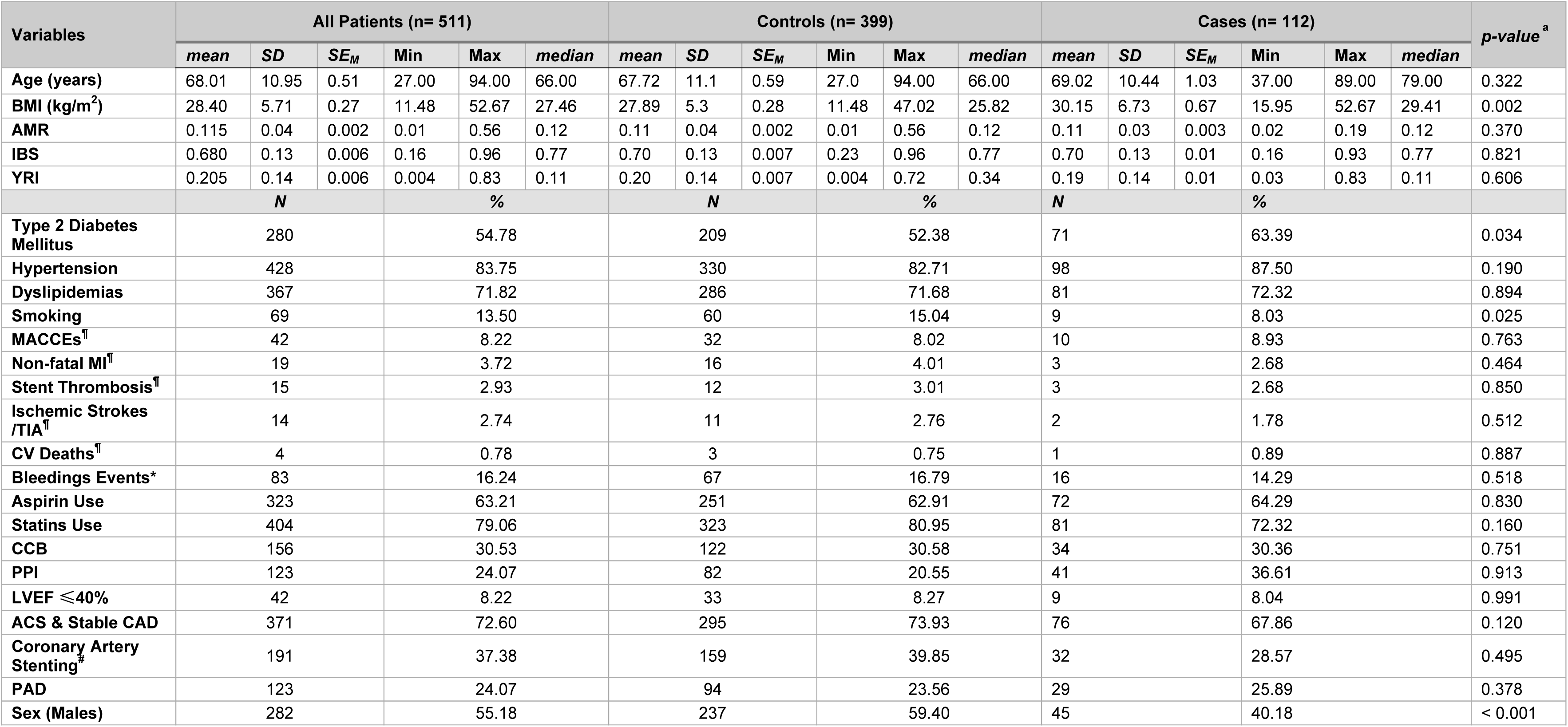
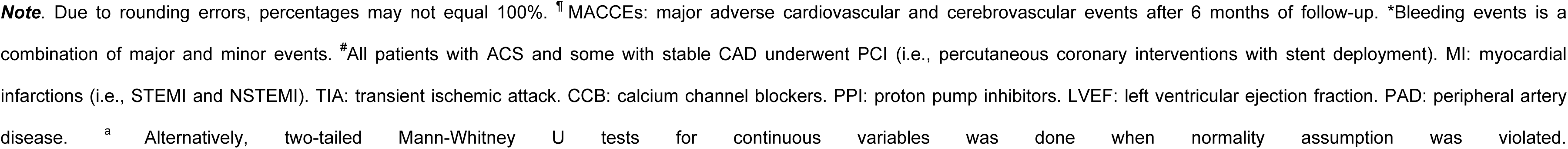
Baseline characteristics of 511 Caribbean Hispanic participants in the study cohort. BMI means body mass index. IBS, AMR and YRI stand for Iberians (European), Native American and Yoruba (African) ancestries, respectively. Significance differences between cases (PRU≥230, n=133) and controls (PRU<230, n=473) were statistically tested by either two-tailed, unpaired *t*-tests for independent samples (continuous variables)^a^ or the two proportions z-tests (nominal variables, %).

| Variables | All Patients (n= 511) |  |  |  |  |  | Controls (n= 399) |  |  |  |  |  | Cases (n= 112) |  |  |  |  |  | p-value<sup>a</sup> |
| --- | --- | --- | --- | --- | --- | --- | --- | --- | --- | --- | --- | --- | --- | --- | --- | --- | --- | --- | --- |
|  | mean | SD | SE<sub>M</sub> | Min | Max | median | mean | SD | SE<sub>M</sub> | Min | Max | median | mean | SD | SE<sub>M</sub> | Min | Max | median |  |
| Age (years) | 68.01 | 10.95 | 0.51 | 27.00 | 94.00 | 66.00 | 67.72 | 11.1 | 0.59 | 27.0 | 94.00 | 66.00 | 69.02 | 10.44 | 1.03 | 37.00 | 89.00 | 79.00 | 0.322 |
| BMI (kg/m<sup>2</sup>) | 28.40 | 5.71 | 0.27 | 11.48 | 52.67 | 27.46 | 27.89 | 5.3 | 0.28 | 11.48 | 47.02 | 25.82 | 30.15 | 6.73 | 0.67 | 15.95 | 52.67 | 29.41 | 0.002 |
| AMR | 0.115 | 0.04 | 0.002 | 0.01 | 0.56 | 0.12 | 0.11 | 0.04 | 0.002 | 0.01 | 0.56 | 0.12 | 0.11 | 0.03 | 0.003 | 0.02 | 0.19 | 0.12 | 0.370 |
| IBS | 0.680 | 0.13 | 0.006 | 0.16 | 0.96 | 0.77 | 0.70 | 0.13 | 0.007 | 0.23 | 0.96 | 0.77 | 0.70 | 0.13 | 0.01 | 0.16 | 0.93 | 0.77 | 0.821 |
| YRI | 0.205 | 0.14 | 0.006 | 0.004 | 0.83 | 0.11 | 0.20 | 0.14 | 0.007 | 0.004 | 0.72 | 0.34 | 0.19 | 0.14 | 0.01 | 0.03 | 0.83 | 0.11 | 0.606 |
|  | N |  |  | % |  |  | N |  |  | % |  |  | N |  |  | % |  |  |  |
| Type 2 Diabetes Mellitus | 280 |  |  | 54.78 |  |  | 209 |  |  | 52.38 |  |  | 71 |  |  | 63.39 |  |  | 0.034 |
| Hypertension | 428 |  |  | 83.75 |  |  | 330 |  |  | 82.71 |  |  | 98 |  |  | 87.50 |  |  | 0.190 |
| Dyslipidemias | 367 |  |  | 71.82 |  |  | 286 |  |  | 71.68 |  |  | 81 |  |  | 72.32 |  |  | 0.894 |
| Smoking | 69 |  |  | 13.50 |  |  | 60 |  |  | 15.04 |  |  | 9 |  |  | 8.03 |  |  | 0.025 |
| MACCEs<sup>¶</sup> | 42 |  |  | 8.22 |  |  | 32 |  |  | 8.02 |  |  | 10 |  |  | 8.93 |  |  | 0.763 |
| Non-fatal MI<sup>¶</sup> | 19 |  |  | 3.72 |  |  | 16 |  |  | 4.01 |  |  | 3 |  |  | 2.68 |  |  | 0.464 |
| Stent Thrombosis<sup>¶</sup> | 15 |  |  | 2.93 |  |  | 12 |  |  | 3.01 |  |  | 3 |  |  | 2.68 |  |  | 0.850 |
| Ischemic Strokes /TIA<sup>¶</sup> | 14 |  |  | 2.74 |  |  | 11 |  |  | 2.76 |  |  | 2 |  |  | 1.78 |  |  | 0.512 |
| CV Deaths<sup>¶</sup> | 4 |  |  | 0.78 |  |  | 3 |  |  | 0.75 |  |  | 1 |  |  | 0.89 |  |  | 0.887 |
| Bleedings Events* | 83 |  |  | 16.24 |  |  | 67 |  |  | 16.79 |  |  | 16 |  |  | 14.29 |  |  | 0.518 |
| Aspirin Use | 323 |  |  | 63.21 |  |  | 251 |  |  | 62.91 |  |  | 72 |  |  | 64.29 |  |  | 0.830 |
| Statins Use | 404 |  |  | 79.06 |  |  | 323 |  |  | 80.95 |  |  | 81 |  |  | 72.32 |  |  | 0.160 |
| CCB | 156 |  |  | 30.53 |  |  | 122 |  |  | 30.58 |  |  | 34 |  |  | 30.36 |  |  | 0.751 |
| PPI | 123 |  |  | 24.07 |  |  | 82 |  |  | 20.55 |  |  | 41 |  |  | 36.61 |  |  | 0.913 |
| LVEF ≤40% | 42 |  |  | 8.22 |  |  | 33 |  |  | 8.27 |  |  | 9 |  |  | 8.04 |  |  | 0.991 |
| ACS & Stable CAD | 371 |  |  | 72.60 |  |  | 295 |  |  | 73.93 |  |  | 76 |  |  | 67.86 |  |  | 0.120 |
| Coronary Artery Stenting<sup>#</sup> | 191 |  |  | 37.38 |  |  | 159 |  |  | 39.85 |  |  | 32 |  |  | 28.57 |  |  | 0.495 |
| PAD | 123 |  |  | 24.07 |  |  | 94 |  |  | 23.56 |  |  | 29 |  |  | 25.89 |  |  | 0.378 |
| Sex (Males) | 282 |  |  | 55.18 |  |  | 237 |  |  | 59.40 |  |  | 45 |  |  | 40.18 |  |  | < 0.001 |

**Note.** Due to rounding errors, percentages may not equal 100%. <sup>¶</sup> MACCEs: major adverse cardiovascular and cerebrovascular events after 6 months of follow-up. \*Bleeding events is a combination of major and minor events. <sup>#</sup>All patients with ACS and some with stable CAD underwent PCI (i.e., percutaneous coronary interventions with stent deployment). MI: myocardial infarctions (i.e., STEMI and NSTEMI). TIA: transient ischemic attack. CCB: calcium channel blockers. PPI: proton pump inhibitors. LVEF: left ventricular ejection fraction. PAD: peripheral artery disease. <sup>a</sup> Alternatively, two-tailed Mann-Whitney U tests for continuous variables was done when normality assumption was violated.

Figure 1 shows the overall and relative distribution of ancestral proportions for the three major ancestral populations (i.e., Native American, European, and West-African) to the trihybrid, admixed genomic makeup of individuals in the study cohort. Four representative LAI plots (RFMix karyograms) from individuals on the extremes and the middle of the Figure 1**-C** (PCA) can be found as a supplemental material (***Figure S2***). Recently admixed populations, like Caribbean Hispanics, have a genome that is reminiscent of a mosaic, with sections inherited from Africa and others inherited from Europe or native/indigenous populations. LA is an important consideration in drug response, particularly in admixed population, at the gene level. Therefore, we performed both traditional GWAS, which were adjusted for genomic PCs, and LAI GWAS, incorporating LA into our association analyses. ***Figure S3*** shows the results of the association tests between PCs and either HTPR or PRU and illustrates the reason for choosing only 2 PCs in further analyses.

We conducted both linear and logistic regression analyses to identify genetic association to PRU and HTPR, respectively. Results from the traditional SNP-based GWAS analyses of PRU and HTPR, adjusted by covariates and PCs, are shown in Figure 2 as both Manhattan and Q-Q plots. Top association hits from these GWAS with the beta (ꞵ) coefficients for the linear (PRU) or Odds Ratios (ORs) for the logistic regressions (HTPR), as well as their corresponding *p*-values and 95% confidence intervals (CI), are presented in **Table 2**. Notably, an overwhelming majority of the lead SNPs (∼99%) were situated within intronic, intergenic or long non-coding RNA (lncRNA)/regulatory regions. Conversely, less than 1% of the variants correspond to exonic variants occurring within coding regions. Although the multivariable linear regression GWAS did not yield any loci with genome-wide significant signals, three loci reached the suggestive significance level of 10^-6^ (Figure 2). In Figure 2, we observed noteworthy associations to PRU levels on chromosomes 3, 14, and 22. Specifically, the variant rs1376606 located in an intergenic region near the *OSBPL10* (Oxysterol Binding Protein Like 10 gene) on chromosome 3p23, exhibited a statistically significant relationship with a reduced PRU value (β: −31.38, 95% CI: −43.66 to −19.09, *p*= 7.75·10^-7^). This SNP also resides within a distal enhancer element and serves as an expression quantitative trait locus (eQTL) for *OSBPL10* in multiple GTEx tissues. In the intronic region of *DERL3* (Derlin-3 gene: degradation in endoplasmic reticulum protein 3) on chromosome 22q11.23, the rs5030613 SNP was associated with a decreased PRU value (β: −33.53, 95% CI: −46.76 to −20.3, *p*= 9.42·10^-7^). On chromosome 14q24.2, rs9323567 within the intronic region of *RGS6* (Regulator of G Protein Signaling 6 gene), was also associated with a decreased PRU value with β of −26.71 (95%CI: −37.27 to −16.15, *p*= 9.85·10^-7^). In the multivariable logistic regression GWAS of the HTPR phenotype, we identified rs116022080 on chromosome 4q22.3 to be associated with an increased risk of HTPR (OR: 3.689, 95%CI: 2.191 to 6.209, *p*= 9.02·10^-7^). This SNP is an intronic variant at the *UNC5C* gene (Unc-5 Netrin Receptor C).

**Figure 2.**
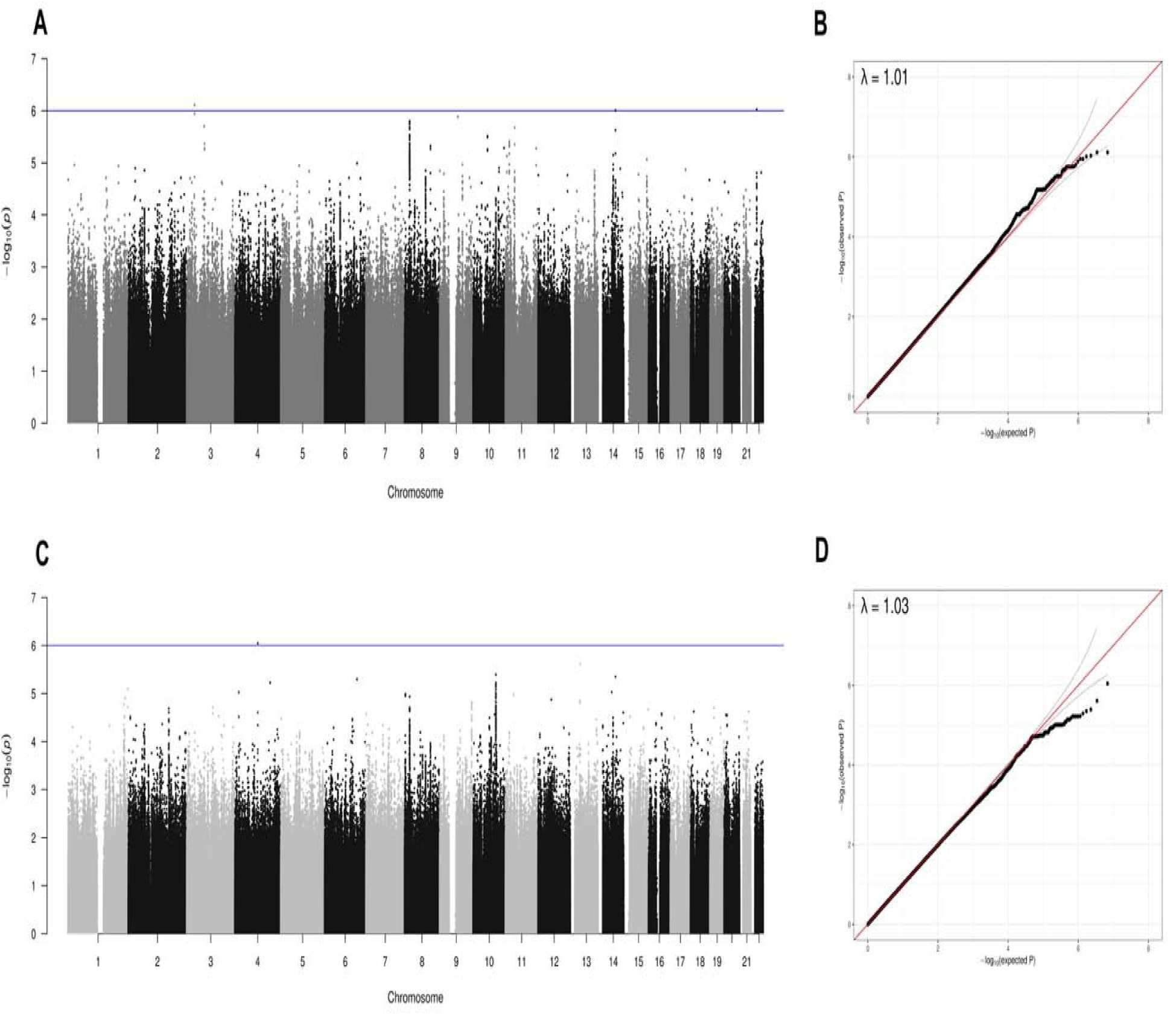
Manhattan and QQ plot from traditional GWAS. Manhattan plot representing the association between single nucleotide polymorphisms (SNPs) and Platelet Reactivity Units (PRU) or High on Treatment Platelet Reactivity (HTPR). The x-axis shows the position of each SNP on the chromosome, while the y-axis displays the association (-log_10_ p-value) between the SNP and the trait of interest. The red line represents the threshold for genome-wide significance (p= 5·10^-8^), and the blue line represents the suggestive level of significance (p= 1·10^-6^). The QQ plot displays the observed p-values of the genome-wide association study (GWAS) summary data on the y-axis, compared to the expected p-values under the null hypothesis of no association on the x-axis. Deviations from the diagonal line indicate departures from the null hypothesis, where points above the line suggest more significant associations than expected, while points below the line suggest fewer significant associations than expected. (A) The GWAS of PRU using linear regression identified 3 loci (*ZNF860* on chromosome 3, *DERL3* on chromosome 22, and *RGS6* on chromosome 14) to be associated with the changes of PRU value at the suggestive level of significance (p< 1·10^-6^). (B) QQ plot for linear regression model GWAS. The genomic control value for the GWAS was λ=1.01, showing no inflation. (C) The GWAS of HTPR using logistic regression showed that one locus on chromosome 4 associated with increased risk of HTPR at the suggestive level of significance (p< 1·10^-6^). (D) QQ plot for logistic regression model GWAS. The genomic control value for the GWAS was λ=1.03, showing no inflation.

**Table 2.**
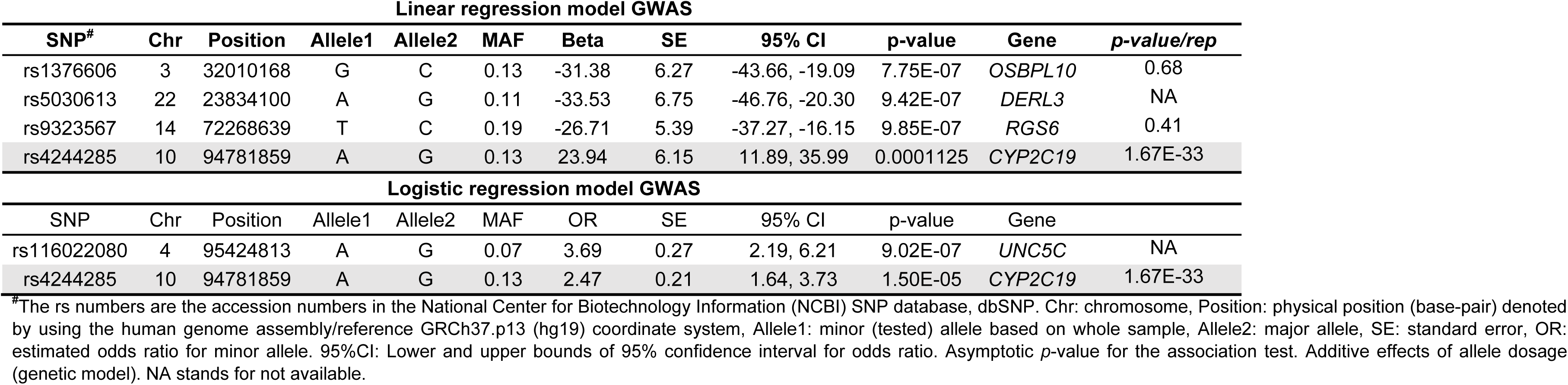
Top SNPs identified by traditional GWAS of platelet reactivity response in 511 Caribbean Hispanics on clopidogrel. Analyses performed in PLINK using both linear and logistic regression and adjusted by age, sex, diabetes, BMI, and PCs. The *p-value/rep* column shows significance of replication in the ICPC cohort (European).

| Linear regression model GWAS |  |  |  |  |  |  |  |  |  |  |  |
| --- | --- | --- | --- | --- | --- | --- | --- | --- | --- | --- | --- |
| SNP# | Chr | Position | Allele1 | Allele2 | MAF | Beta | SE | 95% CI | p-value | Gene | p-value/rep |
| rs1376606 | 3 | 32010168 | G | C | 0.13 | -31.38 | 6.27 | -43.66, -19.09 | 7.75E-07 | OSBPL10 | 0.68 |
| rs5030613 | 22 | 23834100 | A | G | 0.11 | -33.53 | 6.75 | -46.76, -20.30 | 9.42E-07 | DERL3 | NA |
| rs9323567 | 14 | 72268639 | T | C | 0.19 | -26.71 | 5.39 | -37.27, -16.15 | 9.85E-07 | RGS6 | 0.41 |
| rs4244285 | 10 | 94781859 | A | G | 0.13 | 23.94 | 6.15 | 11.89, 35.99 | 0.0001125 | CYP2C19 | 1.67E-33 |
| Logistic regression model GWAS |  |  |  |  |  |  |  |  |  |  |  |
| SNP | Chr | Position | Allele1 | Allele2 | MAF | OR | SE | 95% CI | p-value | Gene |  |
| rs116022080 | 4 | 95424813 | A | G | 0.07 | 3.69 | 0.27 | 2.19, 6.21 | 9.02E-07 | UNC5C | NA |
| rs4244285 | 10 | 94781859 | A | G | 0.13 | 2.47 | 0.21 | 1.64, 3.73 | 1.50E-05 | CYP2C19 | 1.67E-33 |

Of note, the previously validated *CYP2C19*2* (rs4244285) variant was not significant in this study cohort, with p values of 1.5·10^-5^ for HTPR and 1.1·10^-4^ for PRU (**Table 2**). This coding variant had previously been strongly associated with a decreased PRU (p= 1.67·10^-33^) in a large GWAS conducted by the ICPC among individuals of European ancestry.^6^ Results of the LAI adjusted GWAS, conducted to increase the power of detecting association in clopidogrel-treated patients from this admixed cohort and to minimize the impact of local ancestry bias, are depicted in Figure 3. While no variant was genome-wide significant in the ancestry specific analysis (***Figure S4***, A-C), the LAI GWAS meta-analysis identified a near significant association of the intronic SNP rs12571421 (c.819+228A>G), located in the *CYP2C19* gene on chromosome 10, with a higher risk of HTPR (OR: 2.06, 95% CI: 1.54 to 2.74, p= 8.37·10^-7^). Moreover, we found that *CYP2C19*2* (rs4244285) was associated with a higher risk of HTPR (OR: 2.01, 95% CI: 1.51 to 2.31, p= 1.67·10^-6^) in our patients (Figure 3).

**Figure 3.**
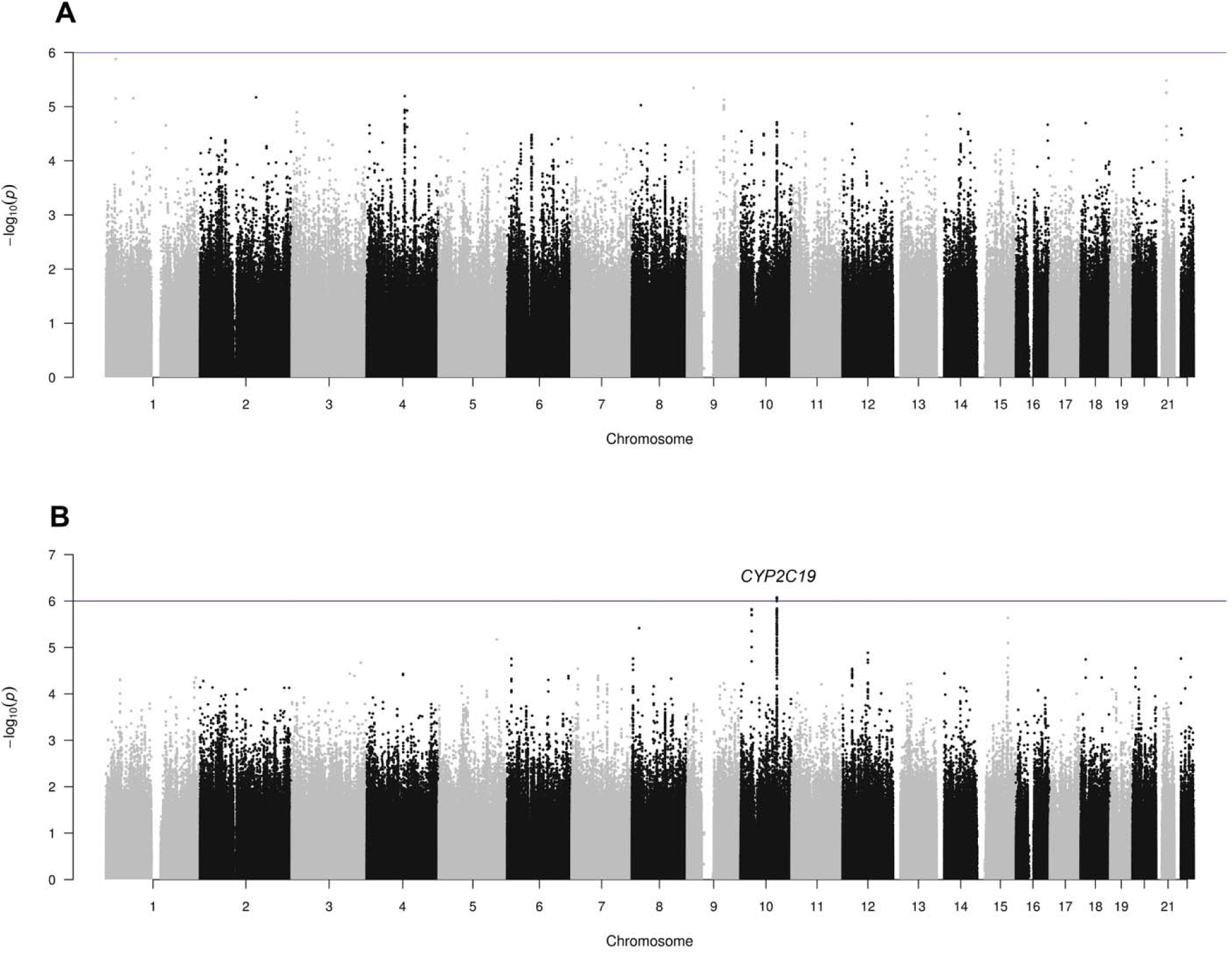
Manhattan plot of LAI adjusted GWAS. After tractor analysis, we used METAL to perform a meta-analysis across all the ancestry to obtain associating results that were adjusted for LAI (A) No variant reached the suggestive significance level in linear regression model local ancestry inference GWAS. (B) The logistic regression model local ancestry inference GWAS showed one locus on chromosome 10 associated with increased risk of HTPR at the suggestive level of significance (p< 1·10^-6^).

Notably, significant effects were observed in individuals of European (p= 0.00088) and African (p= 0.00076) ancestry, but not in those of Native American ancestry (p= 0.26) (**Table 3**). Furthermore, the *CYP2C19*2* (rs4244285) is in strong linkage disequilibrium (LD) with the top SNP hit of this LAI GWAS (r^2^=0.96, D’=1.0 among Hispanics) (Figure 4). The two SNPs are located in an LD block of 6kb, but the intronic SNP was removed from the *CYP2C19*2.003* allele definition by PharmVar due to unknown functionality.^29^ Our findings suggest that *CYP2C19* SNPs may have distinct effect sizes on clopidogrel response based on local genetic ancestry. Importantly, this effect was only seen as suggestive of significance in the LAI adjusted GWAS, suggesting the need for LA adjustment in GWAS of admixed populations such as Caribbean Hispanics. No association was found in the LAI GWAS to PRU. ***Figure S5*** depicts QQ plot to compare the power of regular GWAS and LAI GWAS.

**Figure 4.**
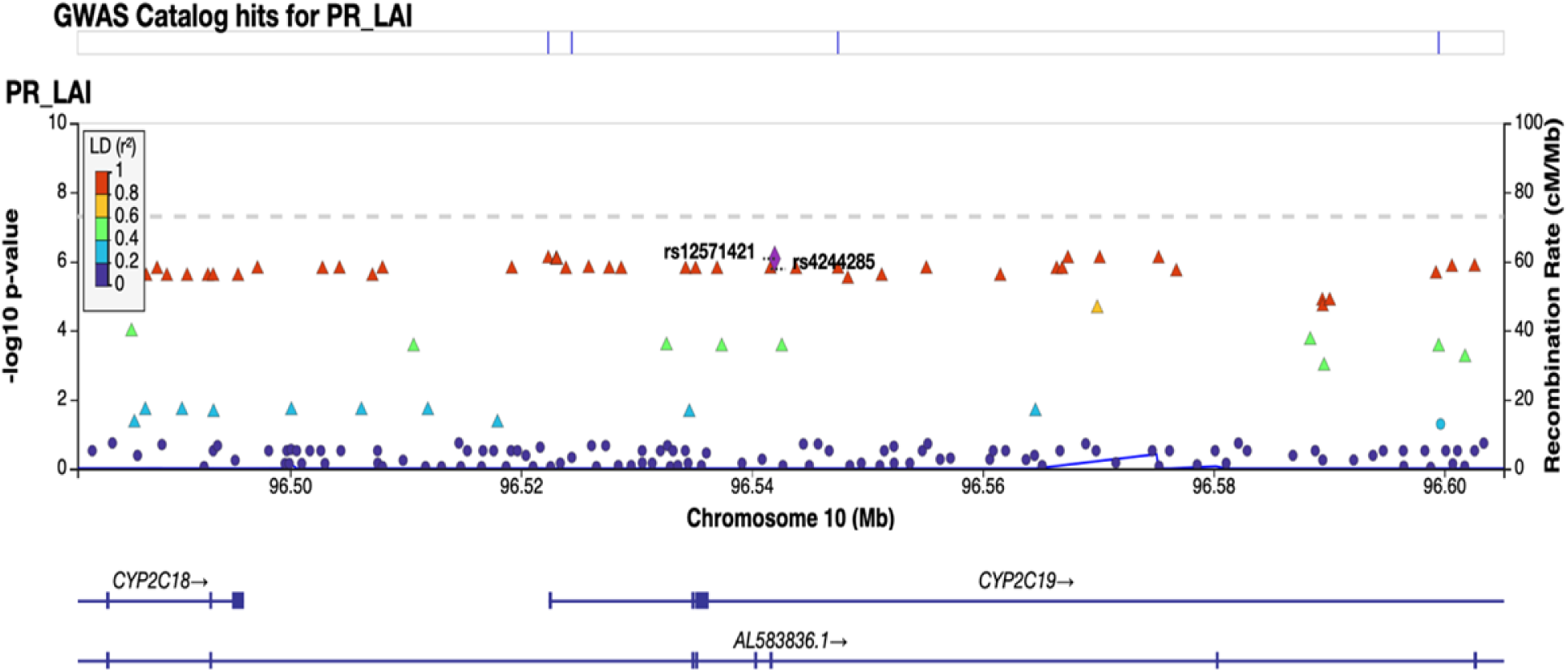
Region plot of top SNP from LAI logistic GWAS. The colors of the circles refer to linkage disequilibrium (LD) (r^2^) between top SNP rs12571421 (purple diamond) and nearby SNPs (based on pairwise r^2^ values from the 1000 Genomes Project reference panel). The blue line and right y-axis show the estimated recombination rate. The x-axis represents the genomic position in chromosome 10 and the left y-axis represents the -log_10_ p-value of association with HTPR in discovery cohort. Region plot showed *CYP2C19*2* variant rs4244285 is high LD with top SNP rs12571421.

**Table 3.**
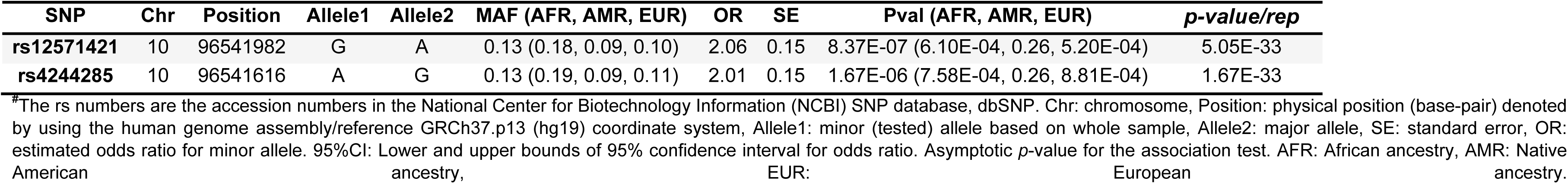
Top SNPs in LAI adjusted GWAS case-control study of HTPR in 511 Caribbean Hispanics on clopidogrel. The *p-value/rep* column shows significance of replication in the ICPC cohort.

The two top signals on chromosome 10 found in our cohort of Caribbean Hispanics from Puerto Rico after performing the LAI-adjusted GWAS driven by the European ancestry/tract were indeed replicated in a previous traditional GWAS of all-European individuals from the ICPC consortium (**Table 3**, rs4244285: p-value of 1.67·10^-33^; rs12571421: p-value of 5.05·10^-33^). However, none of the top hits detected in our traditional GWAS in Caribbean Hispanics following linear and logistic regressions were replicated in the ICPC cohort (**Table 2**) or when tested in either the ACCOuNT consortium cohort of African Americans or the Brazilian cohort (***Figure S6***).

## Discussion

In this study we conducted the first GWAS of clopidogrel response in Caribbean Hispanics from Puerto Rico. Participants in our cohort exhibited a higher average age compared to those in the GWAS conducted by the ICPC focused on platelet reactivity and cardiovascular response to clopidogrel (i.e., 68.01 vs. 64.6 years old, respectively).^6^ However, It is important to highlight that our patient population also displayed a greater degree of genetic admixture and heterogeneity when contrasted with the individuals enrolled in the ICPC study, who were exclusively self-reported as White of European ancestry. Interestingly, although the proportion of males in both studies was greater than females (i.e., 77% in the ICPC cohort and 55% in our cohort), our study exhibited a more balanced distribution of sex. Compared to the ICPC study, a larger proportion of patients from our study had comorbidities that are known to be risk factors for MACCEs. Specifically, the prevalence of type-2 diabetes (55% vs. 24.7%) and hypercholesterolemia (71.8% vs. 64.5%) was higher in our study.^6^ Additionally, the primary indications for clopidogrel therapy were consistent between the two studies, with the majority of participants receiving prescriptions for a combination of ACS and stable CAD (72% vs. 97%). Although our cohort was smaller than the ICPC GWAS, it is the first clopidogrel response GWAS performed with a diverse and previously understudied Hispanic population.

Other than the Native American component, the frequency distributions of ancestral proportions for African and European heritages are broad and shallow and can be interpreted as confirmatory evidence of a diverse genomic architecture, as well as the expected genetic heterogeneity, within Caribbean Hispanics (Figure 1**-A**). Genetically inferred ancestries are not categorical in diverse populations like Caribbean Hispanics, but they rather show a varying degree of admixture proportions (i.e., continuum clines as in Figure 1**-B**) that also scatter in the PCA plot (Figure 1**-C**) with the PCA1 explaining the largest portion of observed variability. Such unique combinations of individual proportions of Native Americans, West-Africans, and Europeans, given by a very distinctive genetic signature in Hispanics, give rise to rich repertoires of allelic combinations and haplotype blocks with very distinctive LD pattern that ultimately affect or even mask true associations in pharmacogenetic studies, becoming an important source of systematic bias. To account for any confounder effect from the variable degree of ancestry on the expected pharmacogenomic associations of clopidogrel resistance in Caribbean Hispanics, we adjusted by PC as a covariate in the regression analyses of regular GWAS and used LAI measures in the Tractor local ancestry GWAS.

Our study is unique in that we used a combination of traditional and LAI adjusted GWAS approaches to identify both novel population-specific genetic regions associated with clopidogrel response and confirm GWAS hits among Caribbean Hispanics from previous studies in Europeans. Importantly, a recent study by Yang et al. identified gene expression signatures associated with increased PRU as well as local ancestry among African Americans in the ACCOuNT consortium cohort and provided new insights into the role of global and local ancestry on clopidogrel response in this underrepresented population.^30^ In our LAI adjusted GWAS meta-analysis, the intronic variant rs12571421 in *CYP2C19*, which is in high LD with *CYP2C19*\*2, was identified as the top signal (OR: 2.06, p= 8.37·10^-7^). In a pharmacogenetic study of clopidogrel in Colombians, *in silico* analysis (SIFT, MutPred, PolyPhen-2) predicted that this intronic SNP is a potential splicing alteration due to activation of a cryptic donor site within *CYP2C19*.^31^ Additional functional studies are needed to determine if the identified association is driven solely by *CYP2C19*2* or a combination of functional genetic effects.

Admixed genomes are made up of portions with varying degrees of distinct ancestries that fluctuate in composition among individuals within the population. These unique kaleidoscopic combinations pose methodological challenges for GWASs in admixed populations like Caribbean Hispanics, including the need to adjust for population stratification. A recent publication assessed the significant impact of cross-ancestry genetic architecture, and the resulting allelic heterogeneity that is given by differences in estimated effect sizes for risk variants across distinct ancestral backgrounds, on GWAS statistics in admixed populations.^32^ By using simulations, authors found that controlling for and conditioning effect sizes on LAI will significantly reduce statistical power.^32^ However, our LAI GWAS in a cohort of Caribbean Hispanics improved power to detect top signal (***Figure S5***) and revealed distinct effect sizes and p-values of relevant SNPs such as *CYP2C19*\*2 on clopidogrel response based on local genetic ancestry. Notably, these signals on chromosome 10 were replicated in the ICPC cohort (**Table 3**). More important, the effect of *CYP2C19*\*2 was strongest in the Europeans tract and weakest in the Native American ancestry suggesting that this allele may be less predictive among individuals from the Caribbean Hispanic population with greater Native American ancestry.

Compared with the traditional GWAS, the QQ plot of LAI GWAS showed the improved power to detect the top signal (***Figure S5***). These findings emphasize the need to account for LAI when conducting a GWAS in highly diverse, admixed populations to control for population stratification and resulting genetic heterogeneity because some significant loci seem to be driven by a particular ancestry group. This is critical and highlights a lack of transferability and poor portability of GWAS findings across diverse populations, particularly in those underrepresented groups with mixed genetic ancestries.

Although no GWAS signals were detected using traditional GWAS methods for PRU and HTPR, a total of 51 loci in 15 different genomic regions including chromosome 10 showed suggestive evidence of association with PRU and HTPR in our study cohort of admixed Caribbean Hispanics (p≤ 5·10^−6^). No significant associations were detected in a previous GWAS in 115 Chinese patients with coronary heart disease (CHD); however, novel variants in chromosomes 9, 18 and 21 that were suggestively associated with antiplatelet effects and clopidogrel pharmacokinetics were identified. Consequently, the predictability of PRU variability at 4 hours in that East Asian population was substantially improved by those novel SNPs despite only showing suggestive evidence of genome-wide association.^33^ In previous GWAS primarily involving individuals of European ancestry, the *CYP2C19* locus on chromosome 10 has consistently demonstrated to be the strongest genetic determinant of the diminished platelet response to clopidogrel treatment, its active metabolite formation, and poorer cardiovascular outcomes.^1–2,6,34^

Despite the modest sample size of the discovery cohort, we were able to identify loci nominally associated with platelet reactivity in Caribbean Hispanics. The effect sizes of these novel loci could likely be a direct consequence of the relatively large percentage of heritability explained by them in this population and the well-defined biological nature of the PRU phenotype. In fact, Shuldiner *et al* have previously established that platelet response to clopidogrel is highly heritable (i.e., h(2)=0.73; p<0.001).^1^ GWAS are generally aimed at finding very small effect sizes and not very low frequency variants, except when such rare SNPs have relatively large effect sizes on the outcome of interest. Accordingly, there are some exceptions to the usually large number of samples needed to confirm small differences with statistical confidence.^35^

The *OSBPL10* gene encodes a member of the OSBP family, a group of intracellular lipid receptors that acts as sterol sensors.^36^ *OSBPL10* has been previously reported as a novel candidate gene for high triglyceride trait in dyslipidemic European patients as well as a regulator of cellular lipid metabolism and apolipoprotein B secretion.^37^A GWAS study in Japanese found that *OSBPL10* SNPs are associated with susceptibility to peripheral arterial disease (PAD).^38^ Dyslipidemia has been largely associated with increased risk of premature CHD and is considered a modifiable factor affecting HTPR in clopidogrel-treated patients.^39–40^ The intergenic variant rs1376606 harbored within a distal enhancer element on chromosome 3 is an eQTL for *OSBPL10* in several GTEx tissues, which may explain the observed association with lower PRUs (*p*= 7.75·10^-7^) in this study.

The intronic rs5030613 variant in *DERL3* was also related to decreased PRU values in our study cohort (*p*= 9.42·10^-7^). A protective role for *DERL3* in the heart has been previously postulated.^41^ The *DERL3* gene encodes a member of the Derlin family of proteins, Derlin-3, which plays a role in ATF6-induced endoplasmic reticulum-associated degradation (ERAD) of unfolded and misfolded glycoproteins to maintain homeostasis under stress.^42^ Derlin-3 overexpression enhanced ERAD and protected cardiomyocytes from ischemia-induced cell death, whereas dominant-negative Derlin-3 increased ischemia-induced cardiomyocyte death. Derlin-3 was also found to be up-regulated in the heart following myocardial infarction (MI), showing cardioprotective effects.^41^

Likewise, an intronic variant within the *RGS6* locus was associated with reduced PRU in our study (rs9323567 SNP; *p*= 9.85·10^-7^). *RGS6* expression is increased in the myocardium of patients with heart disease, and a regulatory role of RGS6 in cardiomyocytes during the aggravation of pathological cardiac hypertrophy has also been reported.^43^ *RGS6* encodes a member of the R7 subfamily of RGS, which is robustly expressed in the heart and is reportedly a key regulator of ischemic injury through its canonical G protein actions. In contrast, the intronic rs116022080 variant in *UNC5C* was associated with an increased HTPR risk in the logistic regression (OR: 3.689, *p*= 9.02·10^-7^). This encoded protein belongs to the UNC-5 family of netrin receptors. At least one *UNC5C* variant was listed among those earlier identified genome-wide significant CAD risk loci, though its role in CAD pathophysiology is still uncertain.^44–45^

Since all the above-mentioned genetic loci have also been found to be associated with high cardiovascular risk, a dual role of these hits as genetic markers of predictive value for both cardiovascular disease susceptibility and clopidogrel resistance cannot be ruled out. However, further studies are warranted to confirm this potential interaction. Despite no individual variant being associated with platelet reactivity at GWAS significance in our cohort of clopidogrel-treated patients, an admixture-adjusted polygenic risk score-driven model using top hits from this study and LAI as weighting factors is expected to help improve predictability of responsiveness to clopidogrel among Caribbean Hispanics. We firmly believe this approach will ultimately facilitate DAPT optimization in this underrepresented population.

The modest sample size of the discovery cohort was a limitation of this study. There is an urgent need to increase representation of diverse populations in pharmacogenomic studies, which has become a high priority for ongoing research initiatives that often build upon existing data. Such a European-centric bias of earlier genetic studies, if not properly mitigated, will limit our understanding of underlying determinants of poor clopidogrel response among Caribbean Hispanics, and will hinder worldwide efforts to adopt a global precision medicine paradigm of true benefit for diverse patients. The current paucity of data from individuals of non-European ancestry due to under-representation in human genetics research will preclude any further extrapolation of derived prediction models to the population at large. Consequently, findings from this work are expected to help in part by addressing such an unmet need and, hence, avoiding the potential harm of extrapolating genetic results from one population to another. This is of remarkable importance given the well-known differences in haplotypes and allele frequency distributions, effect sizes, varying admixture patterns and unique genomic architectures across diverse ethnicities of distinct ancestries.

## Data Availability

The authors confirm that the main data supporting the findings of this study are available within the article and its supplementary materials. The complete data that support the findings of this study will be available in dbGaP at https://www.ncbi.nlm.nih.gov/gap/ (accession number: phs003236.v1.p1), upon request (controlled access).

## Acknowledgements

We would like to thank the patients for voluntarily participating in this study protocol. A special acknowledgement to the Research Design and Biostatistics Core service of *The Alliance*/ Puerto Rico Clinical and Translational Research Consortium (PRCTRC), supported by the NIMHD and the National Institute of Allergy and Infectious Diseases (NIAID) under award # U54MD007587, for helping us with study design and sample size calculations. Additionally, we also want to thank the Bioinformatics Core of the CCRHD-RCMI Program at the UPR-MSC, for their assistance with GWAS analysis, admixture, and ancestry measurements. Special thanks to Edgardo R González, Dr. Ednalise Santiago, Dr. Eduardo Tosado, Dr. Héctor Nuñez, Dr. Ariel González, Dr. Laura Ileana Fernandez-Morales, Dr. Luis Antonio Velez-Figueroa, Dr. Orlando Arce, Frances Marín-Maldonado, Andrés López-Reyes and Marines Rosario for helping us with data collection and patient recruitment. We would also like to thank Dr. Mara Helena Hutz from the Department of Genetics, Instituto de Biociências, Universidade Federal do Rio Grande do Sul, Porto Alegre, Brazil, who kindly provided Brazilian samples from their local biorepository.

## Author contributions

All the authors have accepted responsibility for the entire content of this research article and approved submission. GY, PG, MM, JDS and MAP drafted the manuscript; SAS, GR, ARL, DFHS, KM, MDR assisted with review and editing of the final manuscript; JDS, MAP, SAS, GR, ARL designed the study and help interpreted results; PG, MM, JYR, DFHS, MRB, KM and JD assisted with patient recruitments and data collection; DFHS, KM assessed patient events and adjudicated MACCEs; MM, JYR, MRB, KM, JD performed genotyping and PRU testing; MRB and MDR facilitated access to available databases; GY, PG, MM, KC, CA, JD, MAP performed data analyses, statistical tests and run imputations, QC, GWAS, meta-analysis.

## Disclaimer

The authors are solely responsible for the design and conduct of this study, all study analyses, the drafting and editing of the paper, and its final contents. The contents of this manuscript do not represent the views of the National Institutes of Health (NIH) or the United States Government. No funded writing assistance was utilized in the production of this manuscript.

